# Geospatial disparities in federal COVID-19 test-to-treat program

**DOI:** 10.1101/2022.08.02.22278349

**Authors:** Emily R. Smith, Erin M. Oakley

**Author notes:** Corresponding Author: Emily R. Smith 950 New Hampshire Ave NW, Department of Global Health, Washington, DC, 20052.

## Abstract

**Background:** Paxlovid is authorized for the treatment of COVID-19 and must be used within the first 5 days of symptom onset. This limited window for initiating treatment makes rapid access critical. Federal Test-to-Treat programs provide tests, prescriptions, and medication in one visit^3^.

**Objective:** The objective of this study was to map the location of and identify disparities in access to Test-to-Treat programs in the United States (U.S.).

**Methods:** We obtained location data for public providers of Paxlovid and Test-to-Treat programs in the contiguous U.S. and examined their spatial distribution at the zip code tabulation area level. We defined zip codes as underserved if there was no Test-to-Treat program located within the zip code or within 20 miles of its boundaries.

**Results:** More than 52,000,000 people—representing 16% of the continental U.S. population—do not have access to a Test-to-Treat program in their zip code or within 20 miles. The majority of zip codes representing metropolitan areas have a Test-to-Treat program within 20 miles (77%). In contrast, only 30% of small towns and 23% of rural areas have nearby access. Zip codes with a high proportion of Hispanic and Black residents were likely to have access to nearby Test-to-Treat programs (72%, 70%). In contrast, zip codes with a high proportion of Native American residents were likely to be underserved (70%). About half of high-poverty zip codes do not have access to a Test-to-Treat program within 20 miles.

**Discussion:** Disparities in outcomes related to COVID-19 have been apparent since the beginning of the pandemic and continue to grow. While the multi-dimensional measure of social vulnerability was used to expand the federal Test-to-Treat program, some populations remain without access.

## Introduction

Ritonavir-Boosted Nirmatrelvir (Paxlovid) is an antiviral drug indicated for the treatment of COVID-19 among non-hospitalized adults and children aged ≥12 years who are at high risk of disease progression. In the original trial, Paxlovid reduced the risk of hospitalization or death by 88% among high-risk, unvaccinated adults^1^.

Paxlovid is authorized for use within the first 5 days of symptom onset^2^. This limited window for initiating treatment makes rapid access critical. In March 2022, the Biden-Harris administration established Test-to-Treat programs that provide tests, prescriptions, and medication in one visit^3^. Patients without access to Test-to-Treat programs are advised to contact their primary care provider (PCP) or visit a community health center. However, one in four Americans are without a PCP^4^. In May 2022, additional Test-to-Treat facilities were established to improve access in areas with high social vulnerability^3^. No studies have subsequently examined disparities in access to the program.

The objective of this study was to map the location of and identify disparities in access to Test-to-Treat programs in the United States (U.S.).

## Methods

We obtained location data for public providers of Paxlovid and Test-to-Treat programs in the contiguous U.S. from the Department of Health & Human Services on July 18, 2022. We examined their spatial distribution at the zip code tabulation area level. We defined zip codes as underserved if there was no Test-to-Treat program located within the zip code or within 20 miles of its boundaries. We obtained data on population, urbanicity, race and ethnicity, and poverty for each zip code (see supplemental methods material).

## Results

We found an unequal geographic distribution of Test-to-Treat programs across the United States (Figure 1). More than 52,000,000 people living in 14,812 zip codes—representing 16% of the continental U.S. population—do not have access to a Test-to-Treat program in their zip code or within 20 miles.

**Figure 1.**
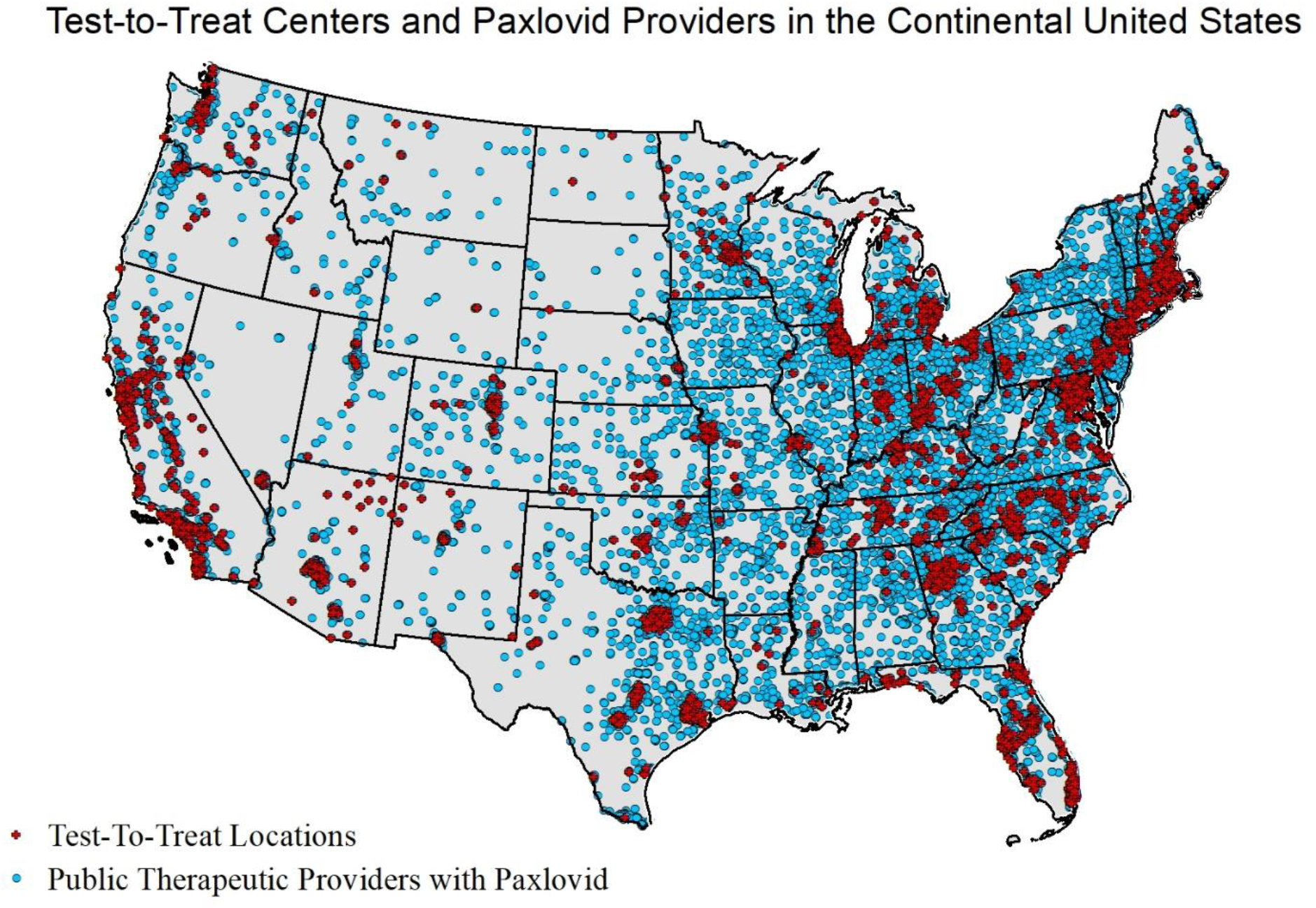
Distribution of Test-to-Treat Centers and Public Therapeutic Providers stocking Paxlovid in the Continental United States, July 18, 2022

The majority of zip codes representing metropolitan areas have a Test-to-Treat program within 20 miles (77%). In contrast, only 30% of small towns and 23% of rural areas have nearby access (Table 1). Zip codes with a high proportion of Hispanic and Black residents were likely to have access to nearby Test-to-Treat programs (72%, 70%). In contrast, zip codes with a high proportion of Native American residents were likely to be underserved (70%). About half of high-poverty zip codes do not have access to a Test-to-Treat program within 20 miles.

**Table 1.** Characteristics of Underserved Zip Codes (Compared to Served) in the Continental United States

|  | **Underserved Zip Codes**<br>(No Test-to-Treat program<br>within 20 miles of zip code)<br>(n=14,812) | **Served Zip Codes**<br>(Test-to-Treat program(s)<br>within 20 miles of zip code)<br>(n=18,488) |
| --- | --- | --- |
| **Urbanicity<sup>a</sup>** |  |  |
| Metropolitan | 4,083 (22.89%) | 13,751 (77.1%) |
| Micropolitan | 3,046 (63.6%) | 1,741 (36.4%) |
| Small town | 2,492 (69.8%) | 1,076 (30.2%) |
| Rural | 4,798 (76.6%) | 1,469 (23.4%) |
| **Race / Ethnicity<sup>b, c</sup>** |  |  |
| Percent Black |  |  |
| High (>37%) | 710 (35.1%) | 1,310 (64.9%) |
| Medium (>2.5-37%) | 2,690 (26.4%) | 7,509 (73.6%) |
| Low (<=2.5%) | 10,900 (54.7%) | 9,041 (45.3%) |
| Percent Hispanic |  |  |
| High (>45.5%) | 449 (28.1%) | 1,149 (71.9%) |
| Medium (>18.3-45.5%) | 896 (27.6%) | 2,349 (72.4%) |
| Low (<= 18.3%) | 12,955 (47.4%) | 14,362 (52.6%) |
| Percent American Indian/Alaska Native |  |  |
| High (>30.1%) | 255 (70.4%) | 107 (29.6%) |
| Medium (>0.7-30.1%) | 9,020 (61.2%) | 5,725 (38.8%) |
| Low (<= 0.7%) | 4,966 (29.3%) | 12,003 (70.7%) |
| **Poverty<sup>b, c</sup>** |  |  |
| High (>17.3%) | 3,974 (49.4%) | 4,069 (50.6%) |
| Medium (>12.3-17.3%) | 2,730 (49.3%) | 2,806 (50.7%) |
| Low (<= 12.3%) | 7,521 (40.9%) | 10,872 (59.1%) |

a. Urbanicity status was drawn from the USDA dataset on rural-urban commuting area codes. Urbanicity status was unavailable for 844 zip codes in the continental United States (including 451 “served” zip codes and 393 “underserved” zip codes).
b. Information on race/ethnicity and poverty status was drawn from the American Community Survey (2020 5-year estimates). Poverty status information was unavailable for 1,328 zip codes in the continental United States (including 741 “served” and 587 “underserved” zip codes). Information on the percentage of Black and Hispanic residents was unavailable for 1,140 zip codes (including 628 “served” and 512 “underserved” zip codes). Information on the percentage of American Indian/Native Alaskan residents was unavailable for 1,224 zip codes (including 653 “served” and 571 “underserved” zip codes).
c. Thresholds for percentage of racial/ethnic subgroups and percent of population in poverty are drawn from the CDC’s COVID Data Tracker, which creates thresholds based on terciles of population characteristics by county.

## Discussion

Disparities in outcomes related to COVID-19 have been apparent since the beginning of the pandemic and continue to grow. We found people living in more rural areas were much less likely to have access to nearby Test-to-Treat programs compared to metropolitan areas. This parallels trends in the cumulative COVID-19 death rate which is increasingly higher among people living in rural and micropolitan areas^5^.

The Test-to-Treat program was specifically designed to reduce barriers to quickly accessing treatment, and indeed nearly half of all sites were located in high vulnerability zip codes by May 2022^6^. However, we found that the majority zip codes with the highest proportion of American Indian and Alaska Native residents were underserved and that almost half of those with a high poverty rate remain underserved. While the multi-dimensional measure of social vulnerability was used to expand the federal Test-to-Treat program, some populations remain without access.

Our findings have several limitations. First, we did not address whether underserved populations successfully access Paxlovid directly through PCPs and community pharmacies. Second, geographic access is only one aspect of ensuring treatment access; previous work found that Paxlovid was under-prescribed in high-vulnerability zip codes despite hosting a high proportion of Test-to-Treat programs^6^.

In an effort to reduce barriers to antivirals, the FDA authorized state-licensed pharmacists to directly prescribe Paxlovid on July 6, 2022^7^. While this is intended to expand treatment access, community pharmacies not participating in the Test-to-Treat program may not offer this service to patients. Public data is needed to understand whether this policy reduces disparities. Further, expanded access must be coupled with increased outreach to patients and providers^6^.

Achieving pharmacoequity is an important goal, especially in the context of an ongoing pandemic with major disparities in health outcomes. Policy efforts, and transparent data, are key to addressing these disparities.

## Data Availability

All data produced in the present study are available upon request to the authors

