## Supplemental Methods Material for "Geospatial disparities in federal COVID-19 test-to-treat program"

Online-Only Material:
Supplemental Methods Material

### Supplemental Methods Material

We obtained 2020 shape files for mapping U.S. states, counties, and Zip Code Tabulation Areas (ZCTA) from the U.S. Census Bureau's TIGER/Line Shapefiles[1].

We obtained shape files used to map COVID-19 Test-to-Treat locations from the U.S. Department of Health & Human Services[2]. We mapped the locations of Test-to-Treat facilities over ZCTA (zip code) boundaries using ArcMap. For each individual Test-to-Treat location, we drew a 20-mile radius around the facility. Using ArcMap, we identified any ZCTAs that fell completely outside the 20-mile radius of all Test-to-Treat facilities and categorized these areas as "underserved" by the Test-to-Treat program.

We obtained shape files used to map the locations of Public Therapeutic Provider locations from the U.S. Department of Health & Human Services[3]. We restricted this data to Paxlovid providers only, and mapped the data over ZCTA (zip code) boundaries in ArcMap.

We gathered data on poverty levels at the ZCTA level using data from the American Community Survey (2020 5-year estimates) via the U.S. Census Bureau's data.census.gov platform (see Table S1701)[4]. Using the poverty cut-offs detailed in the U.S. CDC Covid Data Tracker, we categorized each ZCTA as either "high" (> 17.3%), "medium" (>12.3-17.3%), or "low" (<12.3%) poverty[5]. We merged these data categories with the U.S. Census Bureau ZCTA shapefiles to identify the count and percentage of high, medium, and low-poverty ZCTAs in both "served" and "underserved" areas.

We gathered data on total population and the percentage of residents of Hispanic ethnicity at the ZCTA level using data from the American Community Survey (2020 5-year estimates) via the U.S. Census Bureau's data.census.gov platform (see Table B03002)[6]. We calculated an estimate of percentage of residents by ethnicity from the table estimates for total population and population by Hispanic/non-Hispanic ethnicity. Using the cut-offs for Hispanic/Latino population levels detailed in the U.S. CDC Covid Data Tracker, we categorized each ZCTA as either "high" (> 45.5%), "medium" (>18.3-45.5%), or "low" (≤18.3%) concentration of Hispanic/Latino residents[5]. We merged these data categories with the U.S. Census Bureau ZCTA shapefiles to identify the count and percentage of ZCTA with a high, medium, and low level of Hispanic residents in both "served" and "underserved" areas.

We gathered data on total population and the percentage of Black residents and American Indian/Native Alaskan residents at the ZCTA level using data from the American Community Survey (2020 5-year estimates) via the U.S. Census Bureau's data.census.gov platform (see Table B02001)[7]. We calculated an estimate of percentage of residents by race from the table estimates for total population and population by group. Using the cut-offs detailed in the U.S. CDC Covid Data Tracker, we categorized each ZCTA as having either a "high" (> 37%), "medium" (>2.5-37%), or "low" (≤2.5%) percentage of Black residents[5]. Similarly, we categorized each ZCTA as having either a "high" (>30.1%), "medium" (>0.7-30.01%), or "low" (≤0.7%) percentage of American Indian/Native Alaskan residents[5]. We merged these data categories with the U.S. Census Bureau ZCTA shapefiles to identify the count and percentage of ZCTA with a high, medium, and low level of Black residents and American Indian/Native Alaskan residents in both "served" and "underserved" areas.

To assign an urbanicity category at the ZCTA level, we used data provided by the USDA on rural-urban commuting area codes (RUCAs)[8]. The RUCA coding scheme categorizes zip codes in the U.S. into 10 categories based on urbanicity and local commuting levels. Using the RUCA data, we created a summary variable that categorizes ZCTAs into one of four urbanicity categories (regardless of commuting patterns): metropolitan, micropolitan, small town, and rural. We merged this categorical data with ZCTA shape files to identify the count and percentage of ZCTAs in both "served" and "underserved" areas.

We used cutoff points established by the U.S. CDC's Covid Data Tracker to categorize ZCTAs for demographic groups of interest, including percentage of residents by race/ethnicity (Black, Hispanic, Native American), and percentage of residents below the poverty level. The COVID Data Tracker creates these cutoff points based on terciles of population characteristics by county[5].
